# Resilience factors, pain, and physical activity in adolescent chronic musculoskeletal pain: design and protocol of a pilot phase 2 single-group, non-randomized clinical trial

**DOI:** 10.64898/2026.06.19.26356029

**Authors:** Faith Logan, Madeline Marsh, Adam Hively, Jacqueline Warner, Ann Davis, Jamie L. Jackson, William Black

## Abstract

**Introduction:** Chronic musculoskeletal pain (CMSKP) in adolescence is associated with physical, psychological, social, and academic impairment and increase<u>d</u> risk for chronic pain in adulthood. Although physical activity interventions are an evidence-based approach for managing pediatric chronic pain, many adolescents with CMSKP avoid physical activity due to fear of increased pain, low confidence in physical functioning, and other pain-avoidance behaviors. Resilience-focused interventions targeting self-efficacy, motivation, and mental flexibility may improve engagement in valued activities despite pain. This study describes the design and protocol of the Pain REsilience Promotion for Youth (PREP-Y) intervention, a resilience-focused physical activity intervention for adolescents with CMSKP.

**Methods and analysis:** This single-site, pilot phase 2, single-group, non-randomized clinical trial will enroll 40 adolescents aged 12–17 years with CMSKP from Nationwide Children’s Hospital in Columbus, Ohio, USA. Participants complete questionnaires, objective physical functioning assessments, and physical activity monitoring using activPAL devices as baseline measures. Participants then complete 4 virtual resilience-focused intervention sessions targeting pain resilience, self-efficacy, motivation, and adaptive coping related to physical activity. Garmin watches are used to track activity during the intervention period. Follow-up assessments occur post-intervention and at 3 months post-intervention. Primary outcomes include feasibility and acceptability, assessed through recruitment, retention, attendance, intervention fidelity, and completion of study measures. Exploratory outcomes include physical activity, sedentary behavior, pain-related functioning, pain catastrophizing, kinesiophobia, self-efficacy, and resilience-related constructs.

**Ethics and dissemination:** The study was approved by the Nationwide Children’s Hospital Institutional Review Board. Findings will inform a future randomized clinical trial.

This manuscript reflects protocol version 5.0 dated 23 March 2026.

**Trial registration:** ClinicalTrials.gov: NCT06923891.

**STRENGTHS AND LIMITATIONS OF THIS STUDY:**

- This study evaluates a novel resilience-focused intervention specifically designed to promote physical activity engagement in adolescents with chronic musculoskeletal pain.
- The intervention integrates resilience-building strategies with established pediatric pain management approaches and targets modifiable psychological factors associated with physical activity avoidance.
- The study includes both subjective and objective measures of physical activity and functioning, including wearable activity monitors and standardized physical functioning assessments.
- Safety oversight procedures, adverse event monitoring, and protocol deviation reporting are incorporated given the study’s active enrollment status.
- As a single-group pilot study conducted at a single pediatric institution, the study is not powered to evaluate efficacy or generalizability of clinical outcomes.

## Introduction

Chronic musculoskeletal pain (CMSKP), defined as persistent pain in muscles, ligaments, bones, or joints lasting 3 months or longer, affects up to 38%–40% of adolescents in the United States, with approximately 1 in 3 adolescents reporting reoccurring weekly or monthly CMSKP.^1–5^ Pediatric CMSKP is associated with substantial physical, psychological, social, and academic impairment,^6–14^ and experiencing CMSKP in childhood and adolescence increases the risk of experiencing chronic pain in adulthood.^12,15–18^

Current evidence-based interventions for pediatric chronic pain frequently combine cognitive behavioral therapy (CBT) and physical activity focused approaches to reduce disability and improve functioning.^19,20^ Despite the established benefits of physical activity in managing CMSKP, children and adolescents with CMSKP often avoid physical activity.^21–23^ The Fear Avoidance Model (FAM) provides a useful framework for understanding how chronic pain-related disability develops and persists through activity avoidance.^24–26^ According to the FAM, pain catastrophizing and pain-related fear can lead adolescents to avoid physical and daily activities, which reinforces disability and ongoing pain over time.^27^ Traditional interventions have focused on improving functioning by reducing vulnerability factors identified within the FAM, such as fear and catastrophizing. These interventions may therefore benefit from additional approaches that specifically target resilience processes associated with continued engagement in valued activities while experiencing pain. Resilience-focused approaches may promote functioning by strengthening adaptive factors, such as pain resilience, self-efficacy, motivation, and psychological flexibility.

Phase 1 of this research program evaluated these adaptive factors and their relationship to physical activity engagement among adolescents with chronic widespread musculoskeletal pain.^28^ Preliminary data from Phase 1 identified self-efficacy and motivation as the most impactful intervention targets associated with physical activity engagement. Based on these findings, the current study developed the Pain REsilience Promotion for Youth (PREP-Y) intervention, a resilience-focused program designed to increase physical activity engagement and reduce sedentary behavior in adolescents with CMSKP by specifically targeting self-efficacy and motivation. The conceptual framework that guides the PREP-Y intervention, including fear avoidance and resilience-based pathways related to physical activity engagement, is illustrated in Figure 2.

**Figure 1.**
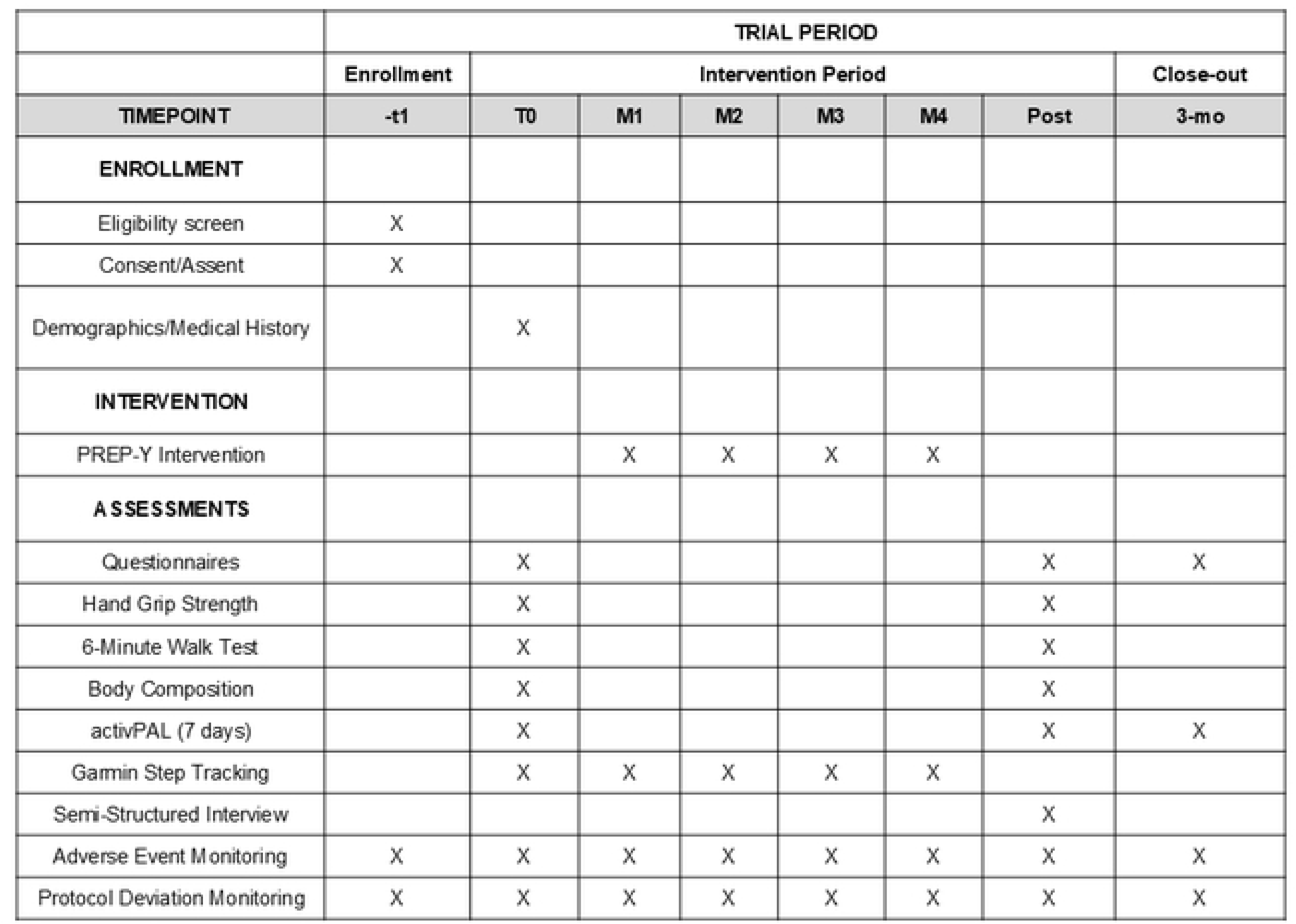
SPIRIT schedule for the PREP-Y trial showing enrollment, intervention delivery, assessments, and follow-up time points. *\* PREP-Y = Pain REsilience Promotion for Youth; activPAL = physical activity monitor; T0 = baseline; T1–T4 = intervention sessions; T5 = post-intervention assessment; T6 = 3-month follow-up assessment*.

**Figure 2.**
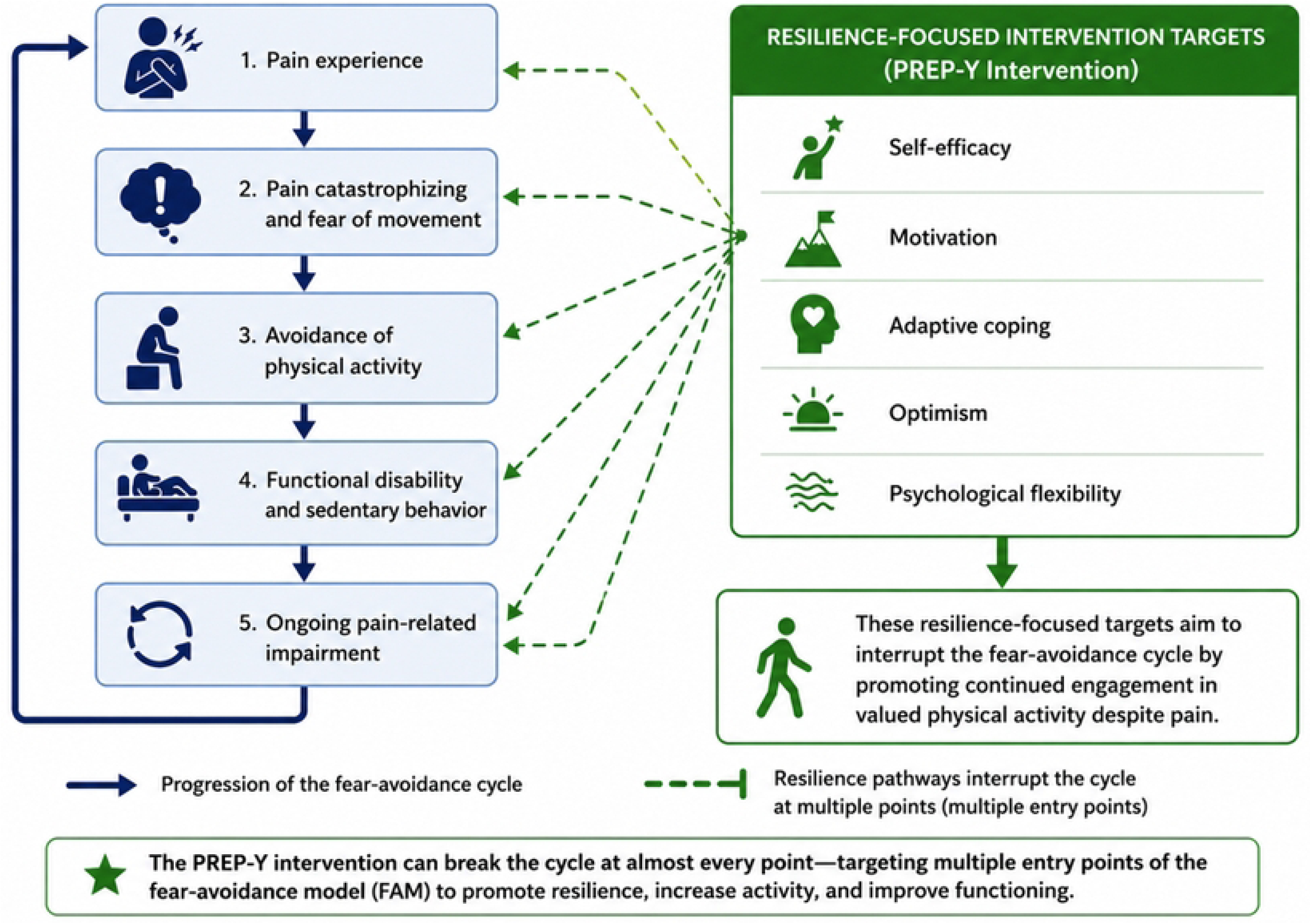
Conceptual framework based on the Fear Avoidance Model and resilience pathway *Resilience-focused intervention targets are hypothesized to interrupt the fear-avoidance cycle by promoting continued engagement in valued physical activity despite pain.

This manuscript presents the design and protocol for an actively enrolling pilot phase 2 clinical trial evaluating the feasibility and acceptability of the PREP-Y intervention in adolescents with CMSKP. Objectives for the pilot phase 2 clinical trial include:

1. The primary objective is to evaluate the feasibility and acceptability of a resilience-focused intervention designed to improve physical activity engagement in adolescents with chronic musculoskeletal pain.
2. The secondary objective is to evaluate the feasibility and acceptability of the physical activity and resilience-related outcome measures and study procedures to inform a future randomized clinical trial.
3. The exploratory objective is to examine preliminary changes in physical activity, sedentary behavior, pain-related functioning, pain resilience, and psychosocial outcomes following participation in the intervention.

## Methods and analysis

### Study design and setting

This study is a pilot phase 2, single-site, single-group, non-randomized clinical trial conducted at Nationwide Children’s Hospital in Columbus, Ohio, USA. The study is designed to evaluate the feasibility and acceptability of a resilience-focused intervention targeting physical activity engagement in adolescents with CMSKP.

Randomization was not used because this study was designed as a pilot phase 2 single-group feasibility and acceptability trial intended to evaluate intervention procedures, participant engagement, recruitment, retention, intervention fidelity, and outcome assessment methods prior to conducting a future randomized clinical trial. A non-randomized design was selected to allow preliminary refinement of the PREP-Y intervention and study procedures before comparative efficacy testing.

Blinding was not performed because participants and study staff were necessarily aware of intervention participation during intervention delivery and assessment procedures. Given the behavioral and interactive nature of the intervention, blinding was not feasible within the context of this pilot feasibility study.

The study began enrollment in July 2025 and is currently actively enrolling participants. Recruitment is anticipated to continue through December 2026, with study completion expected in November 2027. As of June 9, 2026, 198 adolescents had been screened for eligibility, 126 met preliminary eligibility criteria and entered active recruitment, and 27 participants had enrolled in the study with 3 having withdrawn for time constraint issues. 3 participants had completed the full study protocol, and no participants had been lost to follow-up at the time of manuscript preparation. Recruitment and retention metrics continue to be monitored throughout the active enrollment period.

Participants are enrolled for approximately 18 weeks, including baseline assessments, a 4-week intervention period, post-intervention assessments, and a 3-month follow-up assessment. An overview of the study design and participant flow is presented in Figure 3.

**Figure 3.**
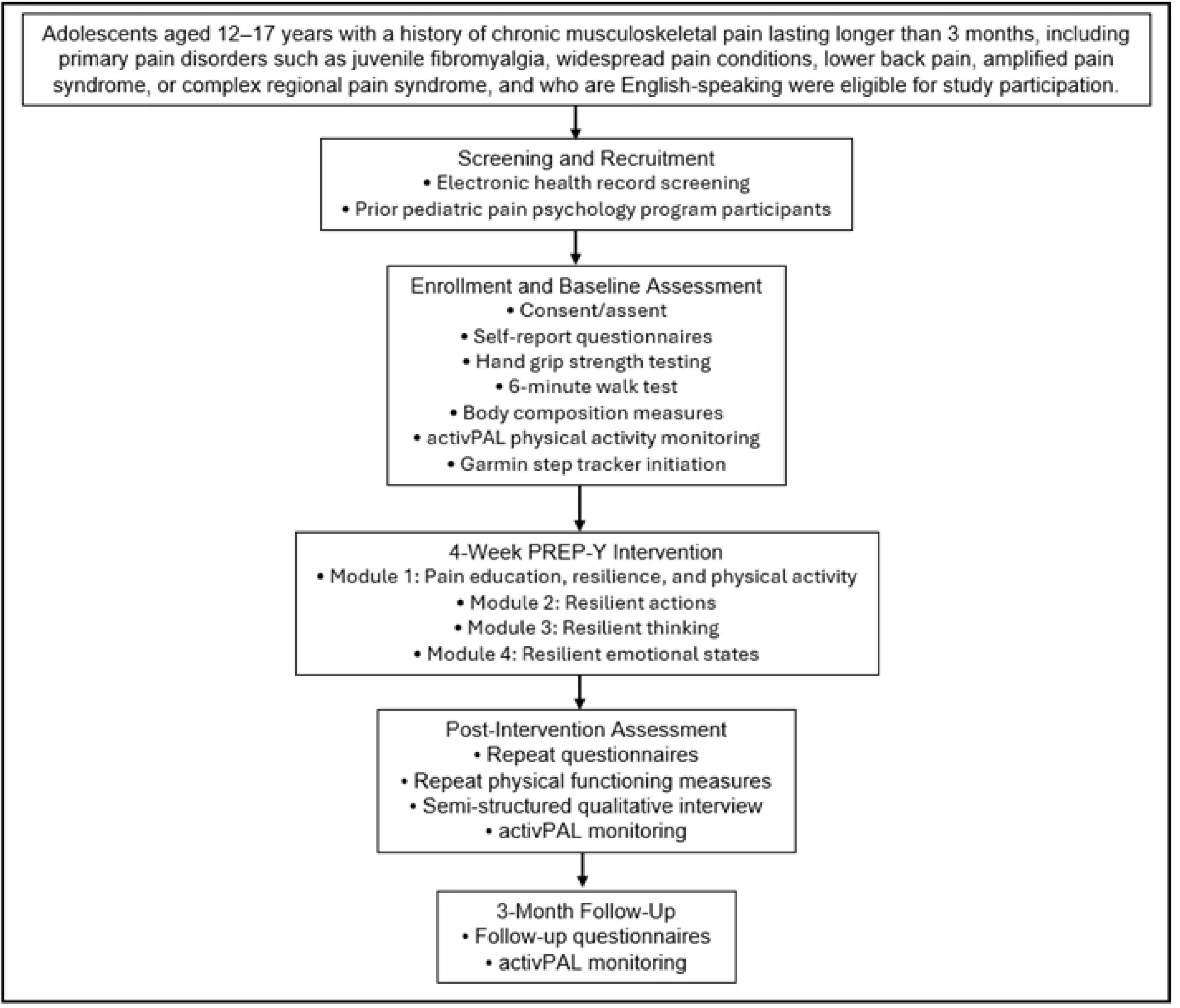
Trial design schematic. This figure illustrates a schematic representation of the pilot phase 2 single-group, non-randomized clinical trial.

### Participants

Participants are recruited from multiple pediatric pain programs at Nationwide Children’s Hospital, including the outpatient Pain Clinic, the Comfort Ability Program (CAP), and the Intensive Pain Rehabilitation Program (iPREP). The outpatient Pain Clinic primarily provides ongoing multidisciplinary outpatient evaluation and management of pediatric chronic pain conditions, CAP is a one-day, structured cognitive-behavioral group intervention for patients and their caregivers focused on pain coping and functioning. iPREP is a 3-week intensive interdisciplinary rehabilitation program designed for adolescents with severe pain-related disability requiring higher levels of rehabilitative care.

Among screened participants, the most common reasons for exclusion included absence of chronic musculoskeletal pain history (n=37), pain secondary to disease (n-15), and age outside study criteria (n=15). After screening, eligible participants and their caregivers receive an opt-out recruitment letter or email prior to direct contact by study staff. If participants do not opt out, study staff conduct follow-up phone contact to assess interest and schedule the initial study visit. The most common non-enrollment reasons among eligible participants included lack of response to recruitment outreach (n=32) and participant decline (n=32).

Recruitment feasibility was informed by prior enrollment patterns within pediatric pain psychology programs at Nationwide Children’s Hospital and the anticipated availability of eligible participants from CAP and outpatient pain clinic referrals.

### Inclusion criteria

1. Adolescents aged 12–17 years.
2. History of chronic musculoskeletal pain for more than 3 months assessed by a pediatric rheumatologist or pain physician.
3. Diagnosis of a primary pain disorder (e.g., juvenile fibromyalgia and other widespread pain conditions, lower back pain, complex regional pain and amplified pain syndrome)
4. Able to read, understand, and speak English.

### Exclusion criteria

1. Chronic pain secondary to disease (e.g., sickle cell disease or rheumatic disease such as juvenile arthritis, systemic lupus erythematosus).
2. Presence of an untreated major psychiatric diagnosis (e.g., major depression, bi-polar disorder, psychoses).
3. History or diagnosis of moderate to severe developmental delay.
4. Patients in foster care or under state guardianship were deemed potentially eligible but excluded, as screening these patients for eligibility was not feasible due to the legal guidelines and restrictions related to screening and enrolling these participants.

### Intervention

Participants completed the PREP-Y intervention during virtual sessions delivered by a trained clinical research coordinator (CRC). The CRC underwent approximately 20 hours of intervention-specific training led by the principal investigators and received ongoing weekly supervision throughout the study to ensure treatment fidelity and adherence to study procedures.

The intervention consists of four weekly individual sessions lasting approximately 60–90 minutes each. A study intervention manual is used to guide discussion, but sessions are individually tailored based on participants’ prior pain education, pain experience, preferred coping approaches, daily routines, and personal interests to increase relevance and engagement. The intervention integrates components from several established pediatric pain and resilience-focused programs while specifically adapting them to target physical activity engagement and movement persistence in adolescents with CMSKP. Components adapted from the FIT Teens intervention include graded activity principles, cognitive-behavioral coping strategies, and approaches promoting functional restoration during pain.^5^ Components informed by previous behavioral interventions for chronic pain include pain neuroscience education, fear-avoidance conceptualization, and strategies supporting activity pacing and daily functioning.^34^ The Resilience Builder Program for Children and Adolescents and the 10 Facets of Highly Resilient People^29^ curriculum also informed intervention content related to optimism, self-efficacy, grit, psychological flexibility, gratitude, and adaptive coping.

Unlike existing pediatric chronic pain interventions that primarily focus on reducing pain-related distress or disability broadly, PREP-Y was specifically developed to target resilience processes associated with physical activity engagement and movement participation despite pain. The intervention therefore emphasizes resilience-based approaches intended to support continued engagement in valued physical and daily activities while addressing motivational barriers, fear-avoidance behaviors, and low confidence in physical functioning commonly observed in adolescents with CMSKP.

#### Module 1

The first module introduces participants to the intervention structure and function and provides education regarding chronic pain, resilience, stress, and physical activity. Participants learn basic pain education principles and thinking strategies for coping with pain.

#### Module 2

The second module focuses on developing proactive strategies for managing pain including problem solving, gratitude practice, and SMART goal setting (Specific, Measurable, Achievable, Relevant, and Time-bound). Participants discuss strategies for addressing physical activity challenges and building adaptive coping patterns.

#### Module 3

The third module emphasizes building cognitive skills for managing chronic pain, including proactive-self-talk, self-compassion, hope and mental flexibility.

#### Module 4

The fourth module introduces self-efficacy, motivation, and grit as resilience processes that support physical activity engagement. Participants practice applying these concepts to current activity-related challenges.

At the beginning of Modules 2-4, participants and interventionists review step count and activity data collected from Garmin activity monitors. Participants and interventionists identify and discuss patterns in physical activity engagement as well as barriers and facilitators to activity participation. This information is then used to inform goal setting, problem solving, and application of resilience-building skills throughout the session.

Between sessions, participants complete brief homework assignments designed to reinforce skill application in daily life. Homework includes practicing coping strategies discussed during sessions (e.g., STOP Sign, self-talk, comfort statements, and proactive coping), setting and tracking SMART goals related to physical activity or daily functioning, completing gratitude or reflection exercises, identifying pain-related thoughts and responses, and applying strategies to real-world situations such as school, work, social activities, or physical activity participation.

A summary of PREP-Y intervention modules, resilience targets, and skill components is provided in Table 1.

**Table 1.** PREP-Y intervention modules.

| Module | Key Topics | Resilience Targets | Skill/Practice |
| --- | --- | --- | --- |
| Module 1 | Pain education, stress, resilience, role of physical activity | Pain resilience awareness | Relaxation strategies, pain education |
| Module 2 | Active coping, optimism, self-compassion | Motivation and adaptive coping | SMART goals, gratitude, STOP technique |
| Module 3 | Cognitive coping and self-talk | Psychological flexibility and adaptive thinking | Positive self-talk, reframing negative thoughts |
| Module 4 | Self-efficacy, motivation, grit | Confidence and persistence with physical activity | Applying resilience skills to physical activity challenges |

\*PREP-Y: Pain Resilience Promotion for Youth. SMART: Specific, Measurable, Achievable, Relevant, Time-bound.

### Outcomes

#### Primary outcomes

Primary outcomes assess feasibility and acceptability of the intervention and study procedures.

Feasibility outcomes include:

- Recruitment rates.
- Retention and study completion rates.
- Session attendance.
- Completion of questionnaires and physical functioning assessments.
- Completion of activity monitoring procedures.
- Intervention fidelity.

Feasibility benchmarks include:

- At least 75% session attendance.
- Intervention delivery fidelity of at least 80%.
- Completion of all study time points by at least 90% of participants.

#### Secondary outcomes: A range of physical activity, functioning, and resilience factors were collected, including

- Physical activity and sedentary behavior measured using activPAL activity monitoring, including step counts, sedentary time, standing time, and physical activity intensity levels (e.g., light activity and moderate-to-vigorous physical activity [MVPA]
- Step counts collected via Garmin devices throughout the intervention period.
- Functional disability measured using the Functional Disability Inventory^30–31^.
- Pain catastrophizing using the Pain Catastrophizing Scale for Children^32^.
- Fear of movement using the Tampa Scale of Kinesiophobia^33–34^.
- Pain intensity measured using visual analogue scales.
- Self-efficacy measured using the PROMIS Pediatric Self-Efficacy Scale^35^.
- Grit measured using the Grit Scale^36^.
- Optimism measured using the Life Orientation Test-Revised^37^.
- Fatigue measured using the PROMIS Pediatric Fatigue Scale^38^.
- Pain coping measured using the PROMIS Pediatric Self-Efficacy Scale^35^.
- Physical functioning measured through hand grip strength testing and 6-minute walk testing.

### Data collection

Data collection occurs at baseline, during the intervention, immediately post-intervention, and at 3-month follow-up (accounting for a two-week window of variability). The schedule of enrollment, intervention delivery, assessments, and follow-up procedures is summarized in Figure 1.

### Baseline assessments

Participants complete self-report questionnaires assessing pain resilience, coping, functioning, self-efficacy (measured using the PROMIS Pediatric Self-Efficacy Scale), catastrophizing (measured using the Pain Catastrophizing Scale for Children (PCS-C)), kinesiophobia (measured using the Tampa Scale of Kinesiophobia), optimism, fatigue (measured using the PROMIS Pediatric Fatigue Scale), and related psychosocial constructs.

Participants also complete objective physical functioning measures including:

- Hand grip strength testing.
- 6-minute walk testing.
- Body composition assessments (height, weight, mid arm circumference, skin trifold thickness).

Participants receive two activity monitoring devices:

1. activPAL monitor for 1-week physical activity assessment.
2. Garmin activity tracker for step monitoring throughout the intervention portion of the study.

The two activity-monitoring devices serve distinct purposes within the study design. ActivPAL devices provide objective assessment of physical activity and sedentary behavior across standardized 1-week monitoring periods at baseline, post-intervention, and follow-up. Garmin devices are used throughout the intervention period to facilitate participant self-monitoring, support session discussions regarding physical activity engagement, and assess intervention adherence and feasibility over time.

### Intervention sessions

Intervention sessions are recorded and transcribed for future qualitative analysis and intervention refinement. Participants also wear the Garmin activity tracker throughout the intervention period. The Garmin-derived step count data are reviewed collaboratively during intervention sessions to support self-monitoring, reinforce goal-setting strategies, facilitate discussion of physical activity barriers, and evaluate feasibility of longitudinal activity tracking procedures.

### Post-intervention assessments

Following completion of the intervention, participants repeat baseline questionnaires and physical functioning assessments. Participants also complete semi-structured qualitative interviews evaluating acceptability of the intervention. Participants reengage in activPAL activity monitoring for 1 week following intervention completion.

### Three-month follow-up

At the 3-month follow-up assessment, participants complete repeat self-report questionnaires and 1 week of activPAL physical activity monitoring to evaluate the maintenance of physical activity, functioning, and resilience-related outcomes following completion of the PREP-Y intervention.

### Safety monitoring and adverse event reporting

Because the study is actively enrolling participants, ongoing safety oversight procedures are in place. Two internally appointed Safety Officers provide safety oversight throughout the study. A formal independent data monitoring committee was not established because this is a single-site, minimal-risk pilot feasibility study evaluating a behavioral intervention without investigational drugs or medical devices. Safety oversight is conducted by internally appointed Safety Officers who review adverse events, serious adverse events, protocol deviations, and study conduct throughout the trial. The Safety Officers are not directly involved in participant recruitment or intervention delivery. Adverse events (AEs), serious adverse events (SAEs), unanticipated problems, and protocol deviations are reviewed and reported in accordance with study procedures and institutional requirements.

### Adverse event definitions

An adverse event is defined as any unfavorable medical occurrence or symptom temporally associated with participation in the study, regardless of whether the event is considered related to the intervention or study procedures.

Potential study-related adverse events include:

- Skin irritation associated with wearable activity monitors.
- Temporary light-headedness following physical functioning assessments such as the 6-minute walk test.
- Pain exacerbations associated with physical activity assessments.
- Emotional distress associated with completion of psychosocial questionnaires.

Serious adverse events are defined as events resulting in death, life-threatening events, hospitalization, or persistent disability.

### Statistical analysis plan

Given the pilot nature of the study, analyses primarily focus on feasibility and descriptive outcomes.

Feasibility and fidelity outcomes, including participant attendance, retention, intervention fidelity, and completion of study measures, will be summarized using descriptive statistics. Session attendance rates will be calculated as the number of completed intervention sessions divided by the total number of scheduled intervention sessions. Study completion rates will be calculated as the proportion of enrolled participants completing all major study time points and outcome assessments. Feasibility benchmarks include at least 75% attendance across intervention sessions, intervention delivery fidelity of at least 80%, and completion of all study time points by at least 90% of participants.

Because this is a pilot feasibility study, analyses will focus primarily on descriptive outcomes rather than formal statistical testing. Exploratory analyses will examine changes in physical activity, sedentary behavior, pain-related functioning, and resilience-related outcomes over time. Mean differences, standard deviations, and preliminary effect sizes will be calculated to help characterize potential changes associated with the intervention and to inform the design of future clinical trials. Findings from these exploratory analyses will be interpreted cautiously and are not intended to establish treatment efficacy.

Missing data patterns will be evaluated descriptively. Given the pilot and feasibility nature of the study, analyses will primarily rely on available data, and missingness will be summarized for all study measures and assessment time points. Activity monitor removal logs, which are used to characterize missing or incomplete activity-monitoring data and study retention data, which is defined as completion of post-intervention and 3-month follow-up assessments among enrolled participants, will also be reviewed to help characterize incomplete physical activity data collection.

The planned sample size of 40 participants was selected to support the primary feasibility and acceptability objectives of this pilot phase 2 study while also providing preliminary estimates of variability and potential intervention effects to inform a future randomized clinical trial (Our overall target enrollment number was increased to 40 participants to accommodate potential participant withdrawals. Our initial goal remains to have 30 participants fully complete the study protocol). An a-priori power analysis was conducted using G*Power, and referenced effect sizes found in the FIT Teens intervention are the basis for PREP-Y.^5,30^ Based upon effect sizes ranging from d = 1.07 to 1.31, as few as n=16 participants may be needed to observe effect sizes at d = 1. The original sample size proposed (n = 30) provides enough power to detect smaller effect sizes (as low as d = .54). With a conservative estimate of 30% drop out, there will still be enough completers (n = 21) for medium to large effect sizes; however, analyses of exploratory clinical outcomes will be interpreted cautiously because the study is not powered to definitively evaluate efficacy.

At the time of manuscript preparation, 26 of the planned 40 participants had enrolled (65.0%). Recruitment proceeded at an average rate of approximately three participants per month, demonstrating strong feasibility of accrual and supporting attainment of the target sample within the projected recruitment timeline.

### Study oversight and data management

The study is conducted at Nationwide Children’s Hospital and led by investigators within the Center for Biobehavioral Health and the Abigail Wexner Research Institute. Weekly research meetings are held to review study progress, participant safety, recruitment, protocol adherence, and study procedures. Data are managed using REDCap electronic data capture systems and stored on secure institutional servers. Detailed data management procedures, including REDCap workflows, quality assurance procedures, fidelity monitoring procedures, and study oversight materials, are maintained by the study team and are available from the corresponding author upon reasonable request. Access to identifiable participant data is restricted to approved study personnel only.

### Patient and public involvement

The intervention was informed by prior research examining resilience, pain coping, and physical activity among adolescents with chronic pain. Existing pediatric chronic pain interventions and resilience-focused treatment frameworks informed intervention development.

Participant feedback collected during post-intervention qualitative interviews will further inform future intervention refinement and development of subsequent clinical trials.

## Data Availability

No datasets were generated or analysed during the current study because this manuscript describes a study protocol. All relevant data generated during the study will be made available upon study completion in accordance with institutional policies, participant consent, and NIH Data Management and Sharing requirements.

## Ethics and dissemination

The study was approved by the Nationwide Children’s Hospital Institutional Review Board. Written parental permission and participant assent are obtained prior to study participation. Participants turning 18 years of age during study participation provide written consent for continued participation.

Important protocol modifications will be submitted for Institutional Review Board approval and updated in the ClinicalTrials.gov registration record, as applicable.

The study includes adolescents with CMSKP and is considered no greater than minimal risk. Participants are informed that they may decline any study procedures or questionnaire items and may withdraw at any time.

Participant confidentiality is maintained through REDCap-based data storage, secure institutional systems, and removal of identifiers following study completion.

The study has a Certificate of Confidentiality through NIH funding mechanisms.

Findings from this pilot trial will be disseminated through conference presentations and peer-reviewed publications and will inform future trials evaluating resilience-focused interventions for physical activity engagement in pediatric chronic pain populations.

## Acknowledgments

The authors acknowledge the contributions of the Behavioral Trials Office staff, research coordinators, participating adolescents and families, and the pediatric pain psychology teams at Nationwide Children’s Hospital.

## Author contributions

FL, MM, MC, and WB contributed to study design and protocol development. WB, JJ, and JW conceptualized the study. FL, MM, and AH coordinated study implementation and regulatory oversight. All authors contributed to manuscript drafting, critically reviewed the manuscript, and approved the final version.

